# Characterization of Dynamic Adaptation to Stressors using Multi-System Stimulus-Response Data: The Study of Physical Resilience in Aging Pilot

**DOI:** 10.1101/2024.12.22.24319519

**Authors:** Karen Bandeen-Roche, Jiafeng Zhu, Qian-Li Xue, Brian Buta, Thomas Laskow, Jeremy D. Walston, Ravi Varadhan

## Abstract

Resilience to stressors has emerged as a major gerontological concept aiming to promote more positive outcomes for older adults. Achieving this aim relies on determining mechanisms underlying capacity to respond resiliently. This paper seeks proof of principle for the hypothesis that physical aspects of said capacity are rooted in the fitness of one’s physiology governing stress response, conceptualized as a dynamical system.

The Study of Physical Resilience in Aging (“SPRING”) leveraged stimulus-response experiments to characterize physiological fitness in older adults scheduled for one of three major stressors: Total knee replacement, incident hemodialysis, or bone marrow transplant in hematological cancer. Here we analyze Holter monitor time series, cortisol responses to adrenocorticotropic hormone (ACTH) stimulation, and repeated diurnal salivary cortisol assessment in the SPRING pilot (n=79). Principal components analysis was applied anticipating steady-state and “adaptation” mechanisms underlying the repeated physiological measures. Analytic features evidenced these mechanisms, supporting construct validity. Component scores were analyzed by major stressor, hypothesized surrogate physiologic measures (physical frailty phenotype, self-report of health), and demographic, health and behavioral characteristics. Scores differed substantially by stressor type and the surrogate physiologic measures, evidencing criterion validity.

Our data support that HRV, ACTH and salivary cortisol stimulus-response data jointly assess adaptation capacity across a variety of major stressors. We believe that SPRING is the first study in humans to concurrently query multiple physiologic systems using stimulus-response tests. Our findings lay groundwork for future validation with further data and to better forecast resilience of older adults to clinical stressors.

## INTRODUCTION

Two adults of comparably late-life age and apparent health experience the same stressor. One soon rebounds, whereas the other is beset by a cascade of adverse events and suffers irreversible decline of health. How can we explain the different responses? What might be done to promote more consistently positive outcomes? These are the hallmark questions of aging. Recent conversations in gerontology have highlighted frailty and resilience as lenses by which to address them. Physical frailty—a state of global vulnerability to stressors (1), and physical resiliencies—ability to rebound from specific stressors (2)—have been theorized to arise in major part from the fitness of one’s physiology governing stress response, energy production, and musculoskeletal integrity (3–6). Henceforth, we term this proposition as the “physiological fitness hypothesis”. The physiology at issue is complex, but its consideration as a multi-component dynamical system might make feasible derivation of a few features largely determining its frailty and resilience (7). If the physiological fitness hypothesis were to be borne out, and then operationalized in clinically relevant measures, this could revolutionize health promotion and care for older adults. Older adults susceptible to planned stressors—such as scheduled clinical procedures—could be identified, and care management be designed accordingly. Long-term tuning of the physiology might forestall precipitous decline, vulnerability, and adverse outcomes to near the end of life.

Such a theory and approach have been well discussed, with many papers evaluating associations of single static physiologic measures with frailty and several evaluating associations of multiple static physiologic measures with frailty at a time (8–10). However, there has been little evaluation of physiologic fitness in humans using dynamic paradigms in which a stimulus is applied and a time series in response is observed, despite that this is the only way that physiology can be studied as a dynamical system (11) and calls to do so were issued decades ago (12,13). One reason why is that such a paradigm is an intensive prospect in studies engaging humans and particularly older adults. The Women’s Health and Aging Studies (WHAS) I and II assessed heart rate variability (HRV) by Holter monitor (14) and 12-hour at-home salivary cortisol trajectories (15)—considering daily life as the provocation; these were performed in distinct subsets (HRV in WHAS I, salivary cortisol in WHAS II). Several years later, WHAS II also implemented a series of stimulus-response tests after its seventh study wave (16–18), finding intriguing but inconsistent hints of relationship to frail status. These studies were limited by small sample size and insufficient overlap in persons receiving various tests, preventing any cross-system analyses. Responding to the sparsity of the published evidence base, the National Institute on Aging (NIA) in 2016 issued a call for research implementing stimulus-response measures to distinguish older adults likely to prove resilient, or not, to forthcoming physical stressors, and identify determinants of these response types. This paper reports findings from a study which arose in response, the Study of Physical Resilience in agING (SPRING; 19).

The SPRING built on the stimulus-response testing begun in the WHAS. SPRING was designed to apply a battery of dynamic stimulus tests in older adults shortly scheduled for planned clinical stressors—total knee replacement, hemodialysis initiation and bone marrow transplant—in order to evaluate (i) the physiological fitness hypothesis with respect to resilience to these stressors, and (ii) the utility of stimulus-response data collection and analysis to accomplish (i). Leveraging data from the SPRING pilot study, this paper seeks to provide, or contradict, proof of principle for the utility of dynamic stimulation data. It will then fall to the primary SPRING study to validate our finding and address (i) above.

In sections to follow, we first further elucidate our idealized conceptual framework as well as a simplification to accommodate data feasibly obtainable in our study setting. We then outline the SPRING study and dynamic stimulus measures and conduct analyses to validate a subset of our dynamic stimulus measures with respect to our conceptual framework. Our report seeks to demonstrate the measures’ potential to reflect underlying physiological system fitness and functioning.

## METHODS

### Conceptual framework

The SPRING conceptual framework is centered on “Physiologic Resilience capacity”— or physiologic fitness as termed in the prior section (19; reproduced for convenience in eFigure 1 in the Supplement). This construct represents “capacity” to respond to a stressor soon to be experienced. As articulated in a previous article, we hypothesize that this capacity is rooted in key inter-connected physiological systems that together maintain bodily homeostasis, including energy regulation systems, the autonomic nervous system, and the innate immune system (3). Also as per that article, we regard the anatomy, functions, and interconnections of these components together as comprising a complex dynamical system— dynamical because the state of the system changes over time, and complex because the component parts interact in “non-simple” ways which generate synergies, evolution, and responses to stimuli that are difficult to infer from the states of the parts (5,20).

Physiologic resilience conceptualized as such cannot be observed directly, but rather must be inferred through indirect assessments—labeled in eFigure 1 as “Dynamic stimulation tests” (topic of the present paper) and “Surrogate measures” (static indicators). In the ideal, complex dynamical systems can be characterized by differential equations governing their interactions and evolution (21). If time-series data on the states-changes just before and following provocation could be collected, the model parameters might be inferred. Given the focus on resilience, such data should establish steady-state, response to a stimulus, and return toward steady-state (7).

Given the moderate sample size in the SPRING pilot and the temporally coarse data that proved feasible to collect while avoiding the imposition of excessive burden on older adults experiencing major life stressors, this paper reports a simplified systems characterization approach. We conceptualized two key features in a stimulus-response setting: the system’s “steady-state” and its “adaptation” capacity. This approach has proven productive in prior studies of physiological system functioning in aging, including the innate immune system (22–23), autonomic nervous system (24), and HPA axis (15).

### Study design

The SPRING study aims to develop a framework by which to identify clinically relevant signatures of resiliency in older adults undergoing physical (clinical) stressors, generate new validated measures by which to identify individuals with impaired capacity for resilience to stressors, and advance a deeper knowledge of the age-related biological changes that impair stress response systems. It seeks ultimately to open the way for the development of novel interventions to improve care management for older adults facing stressful clinical procedures.

The study design has been detailed separately (19). In brief, there was a first, pilot phase to prioritize measures, define phenotypes and refine study protocols followed by a second phase study designed to evaluate the physiological fitness hypothesis, determine predictive utility of dynamic stimulus measures, and study the biology underlying physical resilience. This paper concerns the first phase. In both first and second phases, three clinical stressors were studied: Total knee replacement, dialysis initiation at onset of end stage renal disease, and bone marrow transplant for hematological cancers. Substudies addressing each stressor were designated as the RESilience in TOtal knee Replacement (RESTORE), Resiliency in Dialysis Initiation (ReDI), and REsiliency in older adults UNDergoing BOne marrow transplant (REBOUND) studies. In the pilot phase 20-30 patients were recruited per substudy. At a baseline evaluation scheduled to shortly precede the stressor, they were evaluated by a large battery of measures whose types are outlined in Figure 1: Follow up evaluations primarily of resilience phenotypes and outcomes were implemented at 1-3 months and 4-6 months (one each) depending on the clinical flow for the particular stressor. This paper is focused on the baseline data addressing physiological resilience capacity.

**Figure 1.**
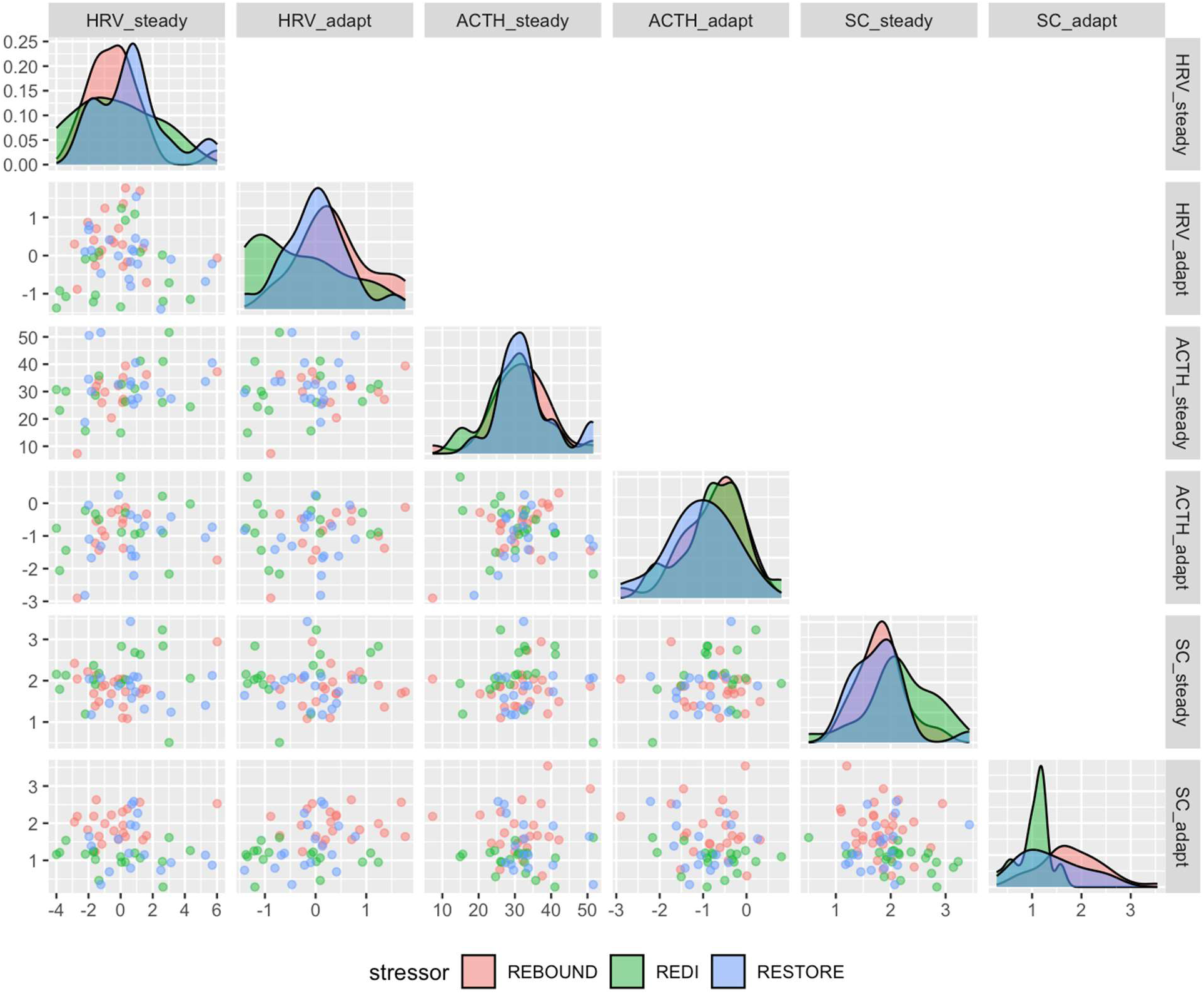
Distributions and co-distributions of physiologic metrics. Distributions are shown as density plots and co-distributions as scatterplots, overlaid by stressor type (identified in the figure key). Shown in order are HRV steady-state and adaptation metrics (fpc1, fpc2), ACTH steady-state and adaptation metrics (cortisol area under the curve and log-log ratio of recovery to baseline), and salivary cortisol steady-state and adaptation metrics (log mean cortisol, log peak-nadir ratio). Alt text: Figure shows density plots comparing steady-state and adaptation physiologic metrics derived from HRV, ACTH and salivary cortisol tests across stressor type, and scatterplots comparing these measures coded for stressor type.

### Dynamic Resilience Capacity Measures

As previously described, SPRING performed seven stimulus-response experiments with participants, designed to query energy metabolism, the hypothalamic pituitary adrenal (HPA) axis, autonomic nervous system (ANS) functioning, and innate immune system activation (19). Only a subset of the resulting measures is available to our analysis due to designed partial deployment of some measures in the first SPRING phase. We focus on three measures assessing the HPA axis and ANS that were widely implemented across all three pilot substudies:

#### Holter ECG monitoring and heart rate variability analysis

During each participant’s baseline assessment, a myPatch Holter monitor was attached to the chest using standardized placement procedures. Patient information and start times were inputted. The recorder’s capacity to mark specific events by tapping on it was used to note the start times of any specific tests during the visit. Each recording stored a 3-channel, digitized ECG signal sampled at 1000 Hz. The monitor was removed at the end of the study visit—usually 2-3 hours after attachment. The stored Holter data were analyzed to research standards (25) by a Holter technician using Cardioscan software developed specifically to interface with the 3-channel myPatch recording. The software detects the onset and morphology of each heartbeat on the recording—a process optimized by the technician based on examination of the ECG strips which display the beat label (*e.g*., N for “normal”, S for “supraventricular”, V for “ventricular”, U for “unclassified or misdetected onset”, or Z for “artifact”) and the time in ms since the last detected “beat.” Optimization includes reanalysis on a different channel or changing the amplitude of a channel. Heart-rate variability (HRV) is calculated from normal-to-normal (N-N) intervals only, hence it is important to accurately identify these intervals (keyed to the R-wave peak). The longest and shortest N-N interval are identified, so that all intervals outside this range are automatically excluded from HRV calculations. A tachogram of instantaneous heart rates, plot of hourly HRV power spectra and scatterplots of each NN interval versus the next are generated to visualize heart rate patterns. Once quality has been assured via the procedures just described, a standard set of HRV values, time domain, frequency domain, and non-linear measures are generated.

#### Adrenocorticotropic hormone (ACTH) stimulation test

This test was performed in the morning in an onsite clinic visit, with or without fasting. Starting times varied between 8 to 10 am. Blood samples were drawn prior to initiation and at 30 and 60 minutes after injection of cosyntropin. Persons taking an oral glucocorticoid were excluded. Plasma cortisol levels were measured using a radioimmunoassay (ALPCO, Salem, NH). The inter- and intra-assay coefficients of variation for cortisol using this assay were 6.30% and 4.65%, respectively.

#### Diurnal salivary cortisol data collection

Participants were sent home from the research unit with a saliva collection kit that included instructions on saliva sample collection and return. Participants were asked to collect saliva at the following times: upon waking/ before eating, 11 a.m., 4 p.m. and just before bedtime, and to record the time of each collection. Salivary cortisol levels were measured using enzyme-linked immunosorbent assay (Salimetrics, Carlsbad, CA). The inter- and intra-assay coefficients of variation for cortisol using this assay were 3.00% and 2.98%, respectively.

### Modeling approach for the dynamic physiologic data

For each of the three tests just described, we defined metrics that we hypothesized to reflect steady-state functioning and adaptation, to be carried forward for further analysis. In each case, “steady-state” refers to the average functioning of the system over the course of the observation time, whereas “adaptation” refers to variation reflecting response to stressors—each in a sense defined further below per test to be examined:

#### HRV steady-state and adaptation

In the WHAS Holter Monitor study outlined above, principal components analysis was applied to summarize key HRV metrics using data from WHAS I and, separately, from the Framingham Heart Study (FHS; 24). We computed two components using the coefficients for the FHS solution. These measures summarize heart rate power in the very low (<0.04 Hz), low (0.04-0.15 Hz), and high (0.15-0.40 Hz) frequency domains as well as standard deviation of N-N intervals (SDNN), root-mean-squared differences of successive N-N intervals (RMSSD), and the proportion of N-N intervals larger than 50 ms (pNN50). The first component (“FPC1”) is essentially an average of the six standardized metrics, reflective of heart rate functioning in steady-state. The second component (“FPC2”) contrasts very low/low with high frequency power as well as SDNN versus RMSSD and pNN50, reflective of adaptive regulation. The reference publication’s authors interpreted higher values of FPC2 as reflective of healthier heart functioning.

#### ACTH steady-state

<u>Area under the curve (AUC)</u> defined by cortisol measurements at times 0, 30 minutes and 60 minutes was calculated by trapezoidal rule. Because the raw cortisol measurements were employed, rather than displacements from the time-0 cortisol measurement, these AUC values were highly correlated with the time-0 cortisol measurement. We opted for the AUC values, which combine 3 measurements together, seeking to reduce measurement error.

#### ACTH adaptation

<u>Recovery</u> was defined as the ratio of the 60-minute measurement to the time-0 measurement. Commensurate with a cortisol surge following cosyntropin administration and then a return to baseline, the ratio of measurements at time t versus time 0 is expected to exceed 1 at times shortly after baseline and then return to a value near 1. Thus, smaller (closer to 1) versus larger values at 60 minutes reflect more complete recovery, and values of 1 indicate complete recovery. This measurement was highly positively skewed: a logarithm-of-logarithm transformation of values succeeded in approximately normalizing it. Log-log values were employed for all statistical analyses.

#### Salivary cortisol steady-state

To represent this, we computed the mean cortisol over the four administration times (in nmol/L). Logarithm values were employed for all statistical analyses: this transformation approximately normalized the distribution.

#### Salivary cortisol adaptation

<u>Peak-nadir ratio</u> was defined as the largest divided by the lowest of the four cortisol measures per person. Logarithm values were employed for all statistical analyses; this transformation approximately normalized the distribution.

### Hypothesized determinants

We sought to explore relationships of physiologic measures to key demographic and health-related determinants we hypothesized to underlie resilience capacity in the initial development of our project. Current age in years was self-reported during screening and verified by the participant during the baseline visit. Sex and race/ethnicity were self-reported, as were smoking history, drinking over the last month, and participation in exercise. Participants were queried as to whether a doctor ever had told them they had each of the following diseases: myocardial infarction, congestive heart failure, angina, chronic pulmonary disease (chronic bronchitis, emphysema, COPD), asthma, liver disease, renal disease, stroke, transient ischemic attack, peripheral neuropathy, hypertension, diabetes, cancer, arthritis, spinal stenosis, osteoporosis, Parkinson’s disease, PAD, venous disease, gastrointestinal ulcer); to balance meaningfulness with adequate counts, number of diseases were dichotomized as 0-2 versus 3 or more. Medication information was collected from the electronic medical record and then reviewed with the participant at a baseline visit to verify accuracy; reported number of medications was dichotomized as 0-4 versus 5 or more. BMI was calculated using height and weight assessed by a trained technician during the study visit.

In addition, we evaluated the associations of the physiologic metrics with the three primary surrogate measures of physiologic resilience capacity hypothesized for the study (eFigure 1 in the Supplement)—frailty, current self-report of health (SRH), and perceived change in health status over the last year. Frailty was assessed in person during a clinic-based study visit according to the physical frailty phenotype as it was implemented in the Women’s Health and Aging Study (26). Self-perception of health (poor, fair, good, very good, excellent), and perceived change in self-reported health over the last year (much better, somewhat better, about the same, somewhat worse, much worse) were assessed via self-report. All three were assessed at the study baseline.

### Data analysis

Study variables were summarized by study, using median (IQR) for continuous variables and percentages for categorical variables. Scatterplots among the physiologic metrics were examined, coded by study.

We then proceeded to analyze the HRV, ACTH stimulation, and salivary cortisol data according to our conceptual framework, pooled across substudies. We implemented correlation-based principal components analysis (PCA) of steady-state and adaptation metrics for the three tests—a total of six metrics. Such an analysis assigns sets of weights—i.e., coefficients—to persons’ multiple *z*-score standardized dynamic test metrics in such a way that the successive weighted combinations of metrics have highest possible variance while being uncorrelated with previous combinations in the set, thus most efficiently distinguish individuals. Following standard terminology, we henceforth label the successive combinations as “components,” their weighted average values as “scores,” and the weights (coefficients) as “loadings.” A frequent goal is to account for shared covariation among metrics being analyzed by only a few components: We hypothesized two components to suffice for our metrics—reflecting systemic steady-state and adaptation capacity. We employed parallel analysis to estimate the number of components reflecting shared variation—a method that employs simulation assuming no shared covariation as a benchmark, and hence avoids overfitting (27). The remaining components are then hypothesized to reflect unshared, measure-specific variation or noise. The sum of the component variances in a PCA matches the sum of the individual variable variances: the ratio of a given component variance to the overall sum is denoted as its “proportion of variance explained.”

As described above, metrics were transformed for approximately normal distribution prior to analysis. Doing so aimed to maximize interpretability of correlations and assist in accommodating missing data (next paragraph). We sought to interpret loading patterns within components as consistent or inconsistent with steady-state and adaptation, hence Varimax rotation was applied to potentially enhance interpretability. This method maximizes loading distinctions within components. We report loadings normalized so that squared values sum to 1: For interpretation, we highlight those loadings whose squared values are higher by a factor of 2 than remaining others as the ones that contribute dominantly to a component score.

Analysis was performed in stages. We first analyzed the complete-case data. Then, considering an appreciable proportion of data missing item-wise in the six continuously scaled metrics, multiple imputation was applied using multivariate normal-based Markov Chain Monte Carlo (MCMC, using Stata function mi imput mvn, version 17 SE; 28) to produce ten dataset replicates filling missing data randomly, using a predictive model. First, datasets were PC-analyzed in parallel. We then synthesized over imputations by performing PCA on the mean HRV, ACTH and salivary cortisol metrics (steady-state, adaptation for each; total of 6 metric means) over replicates. Scores were computed by applying the resulting loadings to the z-transformed metric values.

A final analysis aimed to characterize associations between surrogate measures of physiologic resilience and PC scores, as well as baseline determinants and PC scores. Each score was related to each covariate using linear regression. Crude associations and associations adjusted for SPRING substudy were evaluated. SPRING substudy proved strongly associated with PC scores: We also performed substudy-adjusted analyses restricted to the overlapping range of PC scores represented in all three studies. Scores for the second PC included 3 extreme outliers—one from each substudy: A sensitivity analysis eliminated these from the model adjusting for substudy.

## RESULTS

Seventy-nine older adults participated in the SPRING pilot – 23 in RESTORE, 22 in REDI and 34 in REBOUND (Table 1). Demographic characteristics varied widely between the substudies: RESTORE participants were majority female and majority white; REDI participants, majority male and majority non-white; and REBOUND participants, majority male and nearly all white. Health characteristics also varied, with RESTORE and REDI participants substantially more likely than REBOUND participants to be highly multimorbid, take many medications, and be prefrail or frail. RESTORE participants had considerably higher median BMI and proportions reporting improved health in the last year than those in the other substudies; REDI participants reported the worst current health. Among health behaviors the most striking heterogeneity was observed for exercise, which was endorsed by only 14% of REBOUND participants as compared to more than 40% of RESTORE and REDI participants. In all, the data suggest that the populations undergoing the respective stressors we studied are quite distinct.

**Table 1.** Study of Physical Resilience and Aging Pilot Participant Characteristics by Substudy.

| **Variable** | **RESTORE** | **REDI** | **REBOUND** |
| --- | --- | --- | --- |
| Years of age | 71 (68, 77) | 71 (64, 75) | 67 (65, 72) |
| Female | 16 (70%) | 5 (23%) | 12 (35%) |
| Non-white<sup>a</sup> | 8 (35%) | 14 (64%) | 2 (6%) |
| 3+ Diseases | 17 (77%) | 19 (86%) | 15 (52%) |
| 5+ Medications | 17 (74%) | 19 (86%) | 12 (35%) |
| Body mass index<sup>b</sup> | 32.96 (29.00, 36.13) | 28.60 (24.13, 31.87) | 27.46 (23.72, 30.14) |
| Ever smoking | 8 (36%) | 12 (55%) | 12 (41%) |
| Drank last month | 13 (59%) | 9 (41%) | 14 (52%) |
| Exercise | 10 (48%) | 9 (41%) | 4 (14%) |
| Pre-frail | 14 (64%) | 14 (64%) | 13 (45%) |
| Frail | 3 (14%) | 2 (9%) | 1 (3%) |
| Self-reported health |  |  |  |
| Excellent/Very Good | 7 (32%) | 4 (16%) | 14 (49%) |
| Good | 13 (59%) | 9 (38%) | 12 (41) |
| Fair/Poor | 2 (9%) | 11 (46%) | 3 (10%) |
| Change in self-reported health over last year |  |  |  |
| Much/Somewhat Better | 10 (45%) | 5 (23%) | 6 (21%) |
| Same | 9 (41%) | 13 (59%) | 9 (31%) |
| Somewhat worse/Worse | 3 (14%) | 4 (18%) | 14 (48%) |
| HRV<sup>c</sup> – Steady-state | 0.66 (-0.97, 1.27) | 0.00 (-1.70, 1.22) | -0.41 (-1.52, 0.30) |
| HRV<sup>c</sup> – Adaptation | 0.04 (-0.35, 0.37) | -0.71 (-1.15, 0.09) | 0.30 (-0.07, 0.71) |
| ACTH<sup>d</sup> – Steady-state | 32.32 (27.59, 34.53) | 30.72 (24.67, 33.37) | 32.01 (26.19, 36.71) |
| ACTH<sup>d</sup> - Adaptation | -1.07 (-1.61, -0.47) | -0.76 (-0.95, -0.22) | -0.62 (-1.30, -0.31) |
| Salivary cortisol<sup>e</sup>-<br>Steady-state | 1.81 (1.40, 2.07) | 2.13 (1.91, 2.68) | 1.80 (1.49, 2.04) |
| Salivary cortisol<sup>e</sup>-<br>Adaptation | 1.19 (0.87, 1.93) | 1.13 (0.95, 1.21) | 1.74 (1.46, 2.19) |

*Note.* Abbreviations are: RESTORE-RESilience in Total knee Replacement; REDI-Resiliency in Dialysis Initiation; REBOUND-RESiliency in older adults UNDERgoing BOne marrow transplant; HRV – heart rate variability; ACTH - Adrenocorticotrophic hormone stimulation test.
*Note.* Sample sizes are 23 for RESTORE, 22 for REDI, and 32 for REBOUND. There was incompleteness by selected measures. For numeric measures, data were incomplete for body mass index (1 missing in REDI; 5 missing in REBOUND), HRV (3 missing in RESTORE, 9 missing in REDI, 13 missing in REBOUND), ACTH (5 missing in RESTORE, 7 missing in REDI, 8 missing in REBOUND), and salivary cortisol (4 missing in RESTORE, 4 missing in REDI, 1 missing in REBOUND).
*Note.* Column contents: median (IQR) for numeric measures; n (%) for categorical variables.
a n=21 self-identified as Black; 2, as Asian; 1, as Native American / Pacific Islander. None identified ethnicity as Hispanic.
b Units = kg/m<sup>2</sup>
c Holter monitor data-specific summary scores calculated as per reference #23, Framingham. As PCs, these are unitless.
d Steady-state = area under 0, 30, 60 minute response curve. Adaptation = log(log(60 minute value/time 0 value)). Cortisol was measured as micrograms/dL.
e Steady-state = log(mean of awakening, 11 AM, 3 PM, bedtime measures). Adaptation = log(maximum measure/minimum measure). Cortisol was measured as nmol/L.

The six physiological functioning metrics also are described in Table 1 as well as Figure 1—the latter, presenting scatterplots of the measures against one another as well as distributions stratified by substudy. Focusing on Figure 1: REDI distributions consistently differed from those in the other two studies, most notably comprising the poorest HRV adaptation (lowest FPC2) and salivary cortisol steady-state (highest mean cortisol) values and lacking representation in the better part of the salivary cortisol adaptation (higher peak-nadir ratio) range. Scatterplots demonstrate that transformations primarily succeeded in regularizing the data and, more substantively, associations among the measures were negligible to mild (correlations ranging from −0.25 to 0.30). The Table 1 footnote reports that appreciable data were missing for each measure—typically, 10-30% depending on the study but as high as 40% for HRV in REBOUND.

Principal components analysis was applied to the six (transformed) dynamic measure metrics, beginning with complete-case data (Table 2, Complete-cases column). Two components were rotated: These proved consistent with the steady-state / adaptation theory, with a first component (“PC1”) most highly loaded on two of the adaptation metrics (HRV, salivary cortisol) and a second component (“PC2”) most highly loaded two of the steady-state metrics (HRV, ACTH). Jointly they explained 46% of the variance. Three components, jointly explaining 65% of the variance, had eigenvalues greater than 1: Both component patterns noted above were recapitulated, and an additional component contrasted the salivary cortisol steady-state measure versus the ACTH adaptation measure (eTable 1 in the Supplement; recall that lower ACTH adaptation values reflect better functioning). Despite the encouraging loading patterns and variance explained, the parallel method could not distinguish the correlation pattern observed from an underlying absence of covariation (eFigure 2 in the Supplement). This was not surprising with the small complete-case sample available (n=44), and we proceeded to analyze data multiply imputed 10 times (n=79 per imputed replicate).

**Table 2.** Varimax-rotated Principal Component (PC) loadings of physiologic regulation metrics in Study of Physical Resilience and Aging Pilot.

| Measure | 2 PCs–Complete cases<sup>a</sup> |  | 2 PCs–MI Mean<sup>b</sup> |  | 3 PCs–MI Mean<sup>b</sup> |  |  |
| --- | --- | --- | --- | --- | --- | --- | --- |
|  | PC1 | PC2 | PC1 | PC2 | PC1 | PC2 | PC 3 |
| HRV-Steady-state | -0.08 | 0.56 | -0.02 | 0.55 | -0.09 | 0.72 | -0.03 |
| HRV-Adaptation | 0.60 | 0.33 | 0.53 | 0.34 | 0.75 | 0.05 | 0.20 |
| ACTH-Steady-state | 0.10 | 0.62 | 0.12 | 0.60 | 0.10 | 0.69 | 0.02 |
| ACTH-Adaptation | -0.39 | 0.19 | -0.29 | 0.41 | 0.12 | -0.00 | 0.74 |
| Salivary Cortisol –<br>Steady-state | -0.17 | 0.35 | -0.41 | 0.24 | -0.12 | -0.03 | 0.58 |
| Salivary Cortisol -<br>Adaptation | 0.66 | -0.14 | 0.67 | -0.03 | 0.63 | -0.07 | -0.27 |

*Note.* Abbreviations are HRV – heart rate variability; ACTH - Adrenocorticotrophic hormone stimulation test; PC – principal component; MI – multiple imputation.
a Analysis restricted to cases with all six metrics measured (n=44).
b Loadings derived from mean of each metric (PCA on 6 metric means) over 10 multiple imputations (n=79 per imputation).
c Holter monitor data-specific summary scores calculated as per reference #23, Framingham.
d Steady-state = area under 0, 30, 60 minute response curve. Adaptation = $\log(\log(60 \text{ minute value}/\text{time 0 value}))$ .
e Steady-state = $\log(\text{mean of awakening, 11 AM, 3 PM, bedtime measures})$ . Adaptation = $\log(\text{maximum measure}/\text{minimum measure})$ .

Parallel analysis for PCA of the mean HRV, ACTH and salivary cortisol metrics over replicates suggested 2 components of shared variation (eFigure2). Table 2 reports loadings: Predominant patterns reported for complete-case analysis were recapitulated. Imputation-specific analyses are detailed in the eMethods and in eTable 2 in the Supplement: the number of components identified by parallel analysis varied across replicates, but most frequently this was either two (5 replicates) or three (3 replicates), and in each of these cases, dimensions involving multi-measure steady-state, adaptation, or their contrast were identified. We created PC scores synthesizing replicates as described in the Methods section/Data analysis. Also described in the eMethods in the Supplement is a sensitivity analysis using a second method to create synthetic PC scores, which agreed closely with the primary method.

In addressing potential determinants of physiological functioning, we first contrasted the mean-derived PC distributions by SPRING substudy (Figure 2, top left). Substantial distributional differences were evident, particularly strongly contrasting REDI participants versus the other two substudies. In a linear regression of PC scores on substudy, the mean REDI PC1 score was more than a point lower than in RESTORE (95% CI −1.76 to −0.49) and 1.5 points lower than in REBOUND (95% CI −2.17 to −1.01). Mean differences in PC2 scores were less (smaller than 0.3 points between the maximum, in RESTORE, and the minimum, in REDI, with 95% CIs largely overlapping 0), but REDI scores were strongly bimodal with major mode highest among the studies (Figure 2). We approximated the overlapping ranges, respectively, as −1.5 to 0.18 for PC1 (n=36) and −1.0 to 1.7 for PC2 (n=61). To facilitate interpretation, also shown in Figure 2 are three z-score averages approximating the 3 component solution shown in Table 2: of (i) the salivary cortisol steady-state measure and the ACTH adaptation measure; (ii) the HRV and salivary cortisol adaptation measures, and (iii) the HRV and ACTH steady-state measures. High concordance was observed between (ii) and PC1, and between (iii) and PC2.

**Figure 2.**
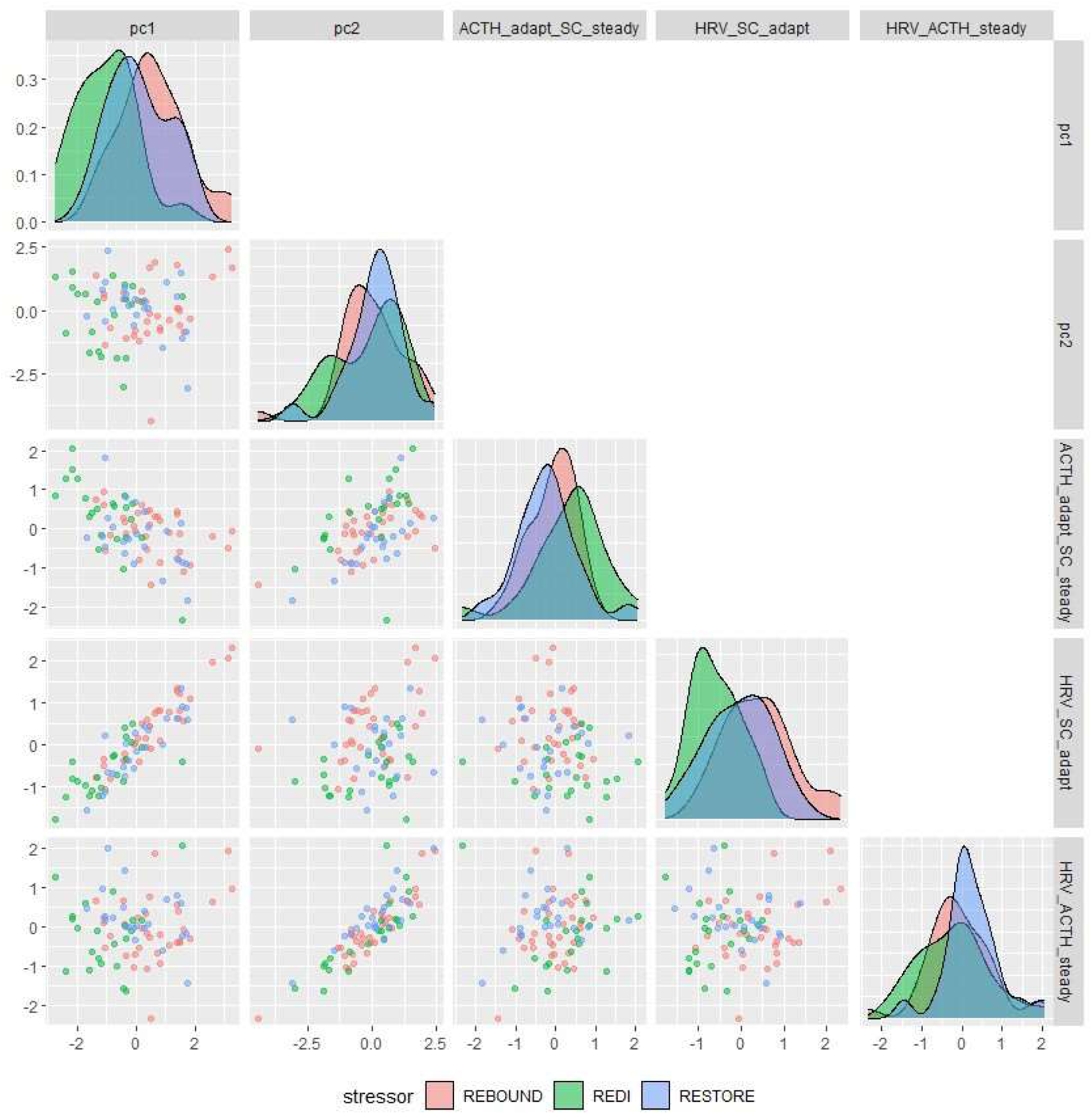
Distributions and Co-distributions of physiological capacity summary measures. Distributions are shown as density plots and co-distributions as scatterplots, overlaid by stressor type (identified in the figure key). Shown in order are multiple imputation mean principal component scores and z-score averages for: ACTH adaptation and salivary cortisol steady-state metrics, HRV and salivary cortisol adaptation measures, and HRV and ACTH steady-state measures. PC1 loads highly on “adaptation” metrics; PC2 loads highly on “steady-state” metrics. Alt text: Figure shows density plots comparing PC and alternative summary measure distributions across stressor type, and scatterplots comparing these measures coded for stressor type. The alternative summary measures are z-score averages for: ACTH adaptation and salivary cortisol steady-state metrics, HRV and salivary cortisol adaptation measures, and HRV and ACTH steady-state measures.

Regressions of PC scores on the hypothesized surrogate measures of physiologic fitness are summarized in Table 3. Robust (as opposed to prefrail or frail) status on the physical frailty phenotype was associated with substantially higher scores on both PCs—crudely, and after adjustment for substudy (in this latter case, an estimated mean difference of 0.61 for PC1=adaptation, 95% CI 0.12 to 1.11, and of 0.60 for PC2=steady-state, 95% CI 0.01 to 1.18). These are shifts of approximately half a standard deviation for each PC. The magnitude of the mean difference was attenuated in the analysis restricting to the range of overlap—this is as expected, given the reduced range of scores in this analysis: Confidence intervals nonetheless remained in the largely positive range (estimated mean difference of 0.33 for PC1, 95% CI −0.03 to 0.68, and of 0.26 for PC2, 95% CI of −0.13 to 0.65). Self-reported health was similarly strongly associated with PC2 scores, with 0.6-1 point mean PC score reductions for good and fair/poor health, respectively, compared to excellent/very good. Fair/poor status was crudely highly associated with reduced PC1 scores, but the association was substantially attenuated after adjustment for substudy. Self-rating of somewhat/much worse health as compared to the previous year showed a similar pattern of association with PC2 scores as did self-reported health, but with smaller estimates of associations and wider confidence intervals, and thus weaker associated evidence.

**Table 3.** Linear Regressions of Multiply Imputed Mean Principal Component (PC) Scores on Surrogate Physiologic Measures, with Stressor Type Adjustment.

| Variable | Model 1 - Crude |  | Model 2 – Stressor<br>type adjustment |  | Model 3 – Stressor<br>type adjustment,<br>overlapping range<sup>a</sup> |  |
| --- | --- | --- | --- | --- | --- | --- |
|  | Coeff | 95% CI | Coeff | 95% CI | Coeff | 95% CI |
| Robust versus<br>prefrail/frail– PC1 | 0.80 | (0.24, 1.36) | 0.61 | (0.12, 1.11) | 0.33 | (-0.03, 0.68) |
| Robust versus<br>prefrail/frail– PC2 | 0.57 | (0.01, 1.14) | 0.60 | (0.01, 1.18) | 0.26 | (-0.13, 0.65) |
| Self-reported health<sup>b</sup> |  |  |  |  |  |  |
| Good – PC1 | -0.07 | (-0.72, 0.58) | 0.16 | (-0.41, 0.74) | 0.15 | (-0.23, 0.52) |
| Good – PC2 | -0.67 | (-1.31, -0.03) | -0.71 | (-1.38, -0.05) | -0.58 | (-1.01, -0.14) |
| Fair/Poor – PC1 | -0.96 | (-1.79, -0.14) | -0.27 | (-1.04, 0.50) | -0.19 | (-0.70, 0.31) |
| Fair/Poor – PC2 | -1.02 | (-1.84, -0.21) | -1.07 | (-1.96, -0.18) | -0.41 | (-1.00, 0.17) |
| Change in self-reported health over<br>last year<sup>b</sup> |  |  |  |  |  |  |
| Same – PC1 | 0.05 | (-0.67, 0.77) | 0.26 | (-0.36, 0.88) | 0.18 | (-0.24, 0.61) |
| Same – PC2 | 0.41 | (-0.27, 1.10) | 0.48 | (-0.23, 1.18) | 0.19 | (-0.30, 0.67) |
| Somewhat/much<br>worse – PC1 | 0.45 | (-0.33, 1.24) | 0.19 | (-0.52, 0.89) | 0.37 | (-0.07, 0.81) |
| Somewhat/much worse – PC2 | -0.56 | (-1.31, 0.19) | -0.63 | (-1.43, 0.17) | -0.24 | (-0.82, 0.34) |

*Note.* Abbreviations are PC – principal component; Coeff – coefficient; CI – confidence interval.
*Note.* Sample sizes are 79 for crude analysis and with stressor type adjustment. See footnote a for sample sizes on restricted ranges.
a Overlapping range is -1.5 to 0.18 for PC1 (n=36) and 1.0 to 1.7 for PC2 (n=61)
b n=6 observations were missing for the self-reported health variables

Few of the other covariates we analyzed showed strong or consistent associations with physiologic functioning PC scores (eTable 3 in the Supplement). The one that endured adjustment for substudy was the association of PC2 with BMI; however, the magnitude of this association was substantially diminished in the sensitivity analysis eliminating three largely outlying points. Associations with robust versus prefrail/frail status and with current self-reported health were unaffected by removal of these outliers; association between PC2 scores and report of somewhat/much worse health over the last year was considerably attenuated.

## DISCUSSION

In this proof of principle study, patterns of correlation among measures of heart rate variability, in-lab adrenocortical stimulation, and at-home diurnal salivary cortisol variation were consistent with theory positing shared physiological governance of these measures’ steady-state levels and adaptation to provocation. In a complete-case data analysis, the large majority of multiply-imputed samples, and a mean of multiple imputations analysis, components predominantly weighted to multiple steady-state measures or to multiple adaptation measures were both evidenced. The great majority of exceptions among the imputations or loadings contrasted steady-state and adaptation. In regression analyses, both PCs were strongly associated with stressor substudy and, more substantively, with robust versus prefrail/frail status after adjusting for stressor substudy. Self-reported health—and, albeit with sensitivity to outliers, BMI—were associated with the steady-state component (PC2) after stressor substudy adjustment. No additional associations with demographic, disease, or behavioral factors were well evidenced.

Our study builds on a considerable body of prior work, while adding important insights. We have noted the WHAS stimulus-response studies: WHAS findings have been separately reported from repeated salivary cortisol assessment at home over 12 hours (15), Holter monitor assessment during a 2-3 hour at-home study evaluation (14), an ACTH test (16), an OGTT (17), and an evaluation of phosphocreatine recovery in response to exercise (18). These are consistent with our findings of association with physical frailty phenotype status. The Baltimore Longitudinal Study on Aging (BLSA) and The Irish Longitudinal Study on Ageing have assessed orthostatic blood pressure in its participants (29–30); the BLSA and various other epidemiological cohorts on aging have conducted oral glucose tolerance tests on their study cohorts (e.g., 31-32). We are not aware of any prior studies that have analyzed data from multiple stimulus-response tests that aimed to query differing physiologic systems concurrently, however. The studies cited earlier in this paragraph, moreover, were observational. We believe that our study and the PRIME-KNEE Study (33) are the first to have designed multiple-stimulus-response test batteries explicitly to characterize resilience capacity in older adults approaching an impending clinical stressor.

Hypotheses positing that multi-system dysregulation underlies frailty have appeared in the literature for at least 20 years (e.g. 1,5,7,8,34,35), and a more recent literature has posited the same for resilience (3,36). We believe that our study is the first to have evidenced these using dynamic, multi-system data. That self-reported health also evidences association with multi-system dysregulation, as SPRING hypothesized during its design, adds proof of principle that the data we are collecting and the steady-state/adaptation paradigm we have applied to summarize it embody meaningful physiological signal. We look forward to validating our findings in the larger sample—allowing for more sophisticated analyses and fuller adjustment for potential confounders—that the primary SPRING study will provide. The primary SPRING hypothesis that physiologic measures are associated with resilient/non-resilient trajectories following a subsequent clinical stressor additionally will be tested in the main study.

We observed substantial heterogeneity in our physiological functioning metrics and PC scores across clinical stressor groups. This adds concurrent validity to the measures we have developed—older adults with late-stage chronic kidney disease predictably have more dysregulated physiology than older adults approaching an elective total knee replacement, for example. To have recruited older adults with a wide range of physiological fitness is a benefit for SPRING. The substantial inter-study heterogeneity, however, also complicates the interpretation of findings. Stressor subgroup was strongly associated with key demographic determinants in the SPRING pilot, including age, sex, race, and BMI. Where substantial crude associations attenuated greatly upon adjustment for substudy, we consider it most probable that the variable was acting as a proxy for substudy (e.g. for nonwhite race, whose representation in REDI was much greater than in the other studies). Where they persisted despite such adjustment but were considerably attenuated in the sensitivity analysis (e.g., BMI), we will look to the primary study to provide further clarity.

### Strengths and limitations

Our study’s overarching strength was its collection of multiple stimulus-response measures in a cohort of older adults approaching major clinical stressors, using rigorous protocols. This is a major innovation. Primary limitations are our relatively small sample size, commensurate with an early-phase pilot study, and frequently missing data on one or more stimulus-response tests. We implemented multiple imputation to mitigate the missing data concern: This approach yields valid estimates of the covariation underlying PCs so long as the data are missing at random, meaning that missingness did not differ by unobserved physiologic status after accounting for the information observed. We believe this assumption is reasonable, given that missed tests often occurred due to scheduling difficulties. The sample size mandated simplifications in analyses, including that few statistical adjustments could be made while avoiding overfitting. We ideally would have fit differential equations to our stimulus-response data, or at least, latent variable analyses hypothesizing “steady-state” and “adaptation” factors. Instead, we evaluated “steady-state” and “adaptation” metrics of the multi-system response using PCA. PCA approximates a latent variable fit, hence provides sound groundwork for future validation. We do not claim that the steady-state/adaptation schema or the stimulus-response tests analyzed in this paper are ideal The SPRING ACTH data collection lasted only one hour: Longer duration would have allowed for more complete cortisol recovery. The salivary cortisol loading’s positive sign in the steady-state PC surprised us: We would have expected negative sign considering elevated daily cortisol as a sign of adverse health. Future validation will be particularly important here. We are reassured that the HRV and ACTH metrics most strongly anchored this PC. Of note, confidence intervals reported in Table 3 do not incorporate imputation-associated uncertainty in principal components loadings. This will be important to do in an ultimate confirmatory study.

A final limitation is the difficulty in recruiting older adults exhibiting the whole range of physiologic fitness in a study like SPRING. Conducting stimulus-response tests is challenging even under ideal conditions, but it is substantially harder when older adults are preparing to undergo major clinical stressors to treat their underlying health issues (37). As we have previously reported, it seems likely that our sample is biased toward more resilient individuals who were willing to undergo the study battery we designed (19). Our data suggest that persons exhibiting a heterogeneous physiologic range nonetheless were recruited—but efforts to facilitate frail and non-resilient individuals in participating in studies such as ours will continue to be important.

Our study supports the usefulness of physiologic steady-state/adaptation as an organizing principle for the summary of dynamic stimulation data spanning HRV, ACTH and salivary cortisol tests. It also evidences that steady-state and adaptation measures summarize physiological functioning relevant to frailty and hence resilience. Our future work will seek to validate this schema with more extensive data, additional dynamic stimulation measures, and more sophisticated modeling, and to evaluate resulting measures’ utility for distinguishing individuals who prove resilient, or not, following the clinical stressors we are studying. We hypothesize that the relevance of the concepts studied here generalizes to other tests, physiological systems, and stressor types.

## Acknowledgments

Assays for plasma and salivary cortisol were performed by the Research Laboratory Core of the Institute for Clinical and Translational Research at Johns Hopkins University.

## Data availability

The SPRING is ongoing. Data will be shared according to the NIH-approved sharing plan for the study; in short, data requests will be reviewed on a per-case basis once primary study analyses have been completed.

## Preprint status

This work has not yet been peer reviewed.

## Supplementary Information

**eFigure 1.**
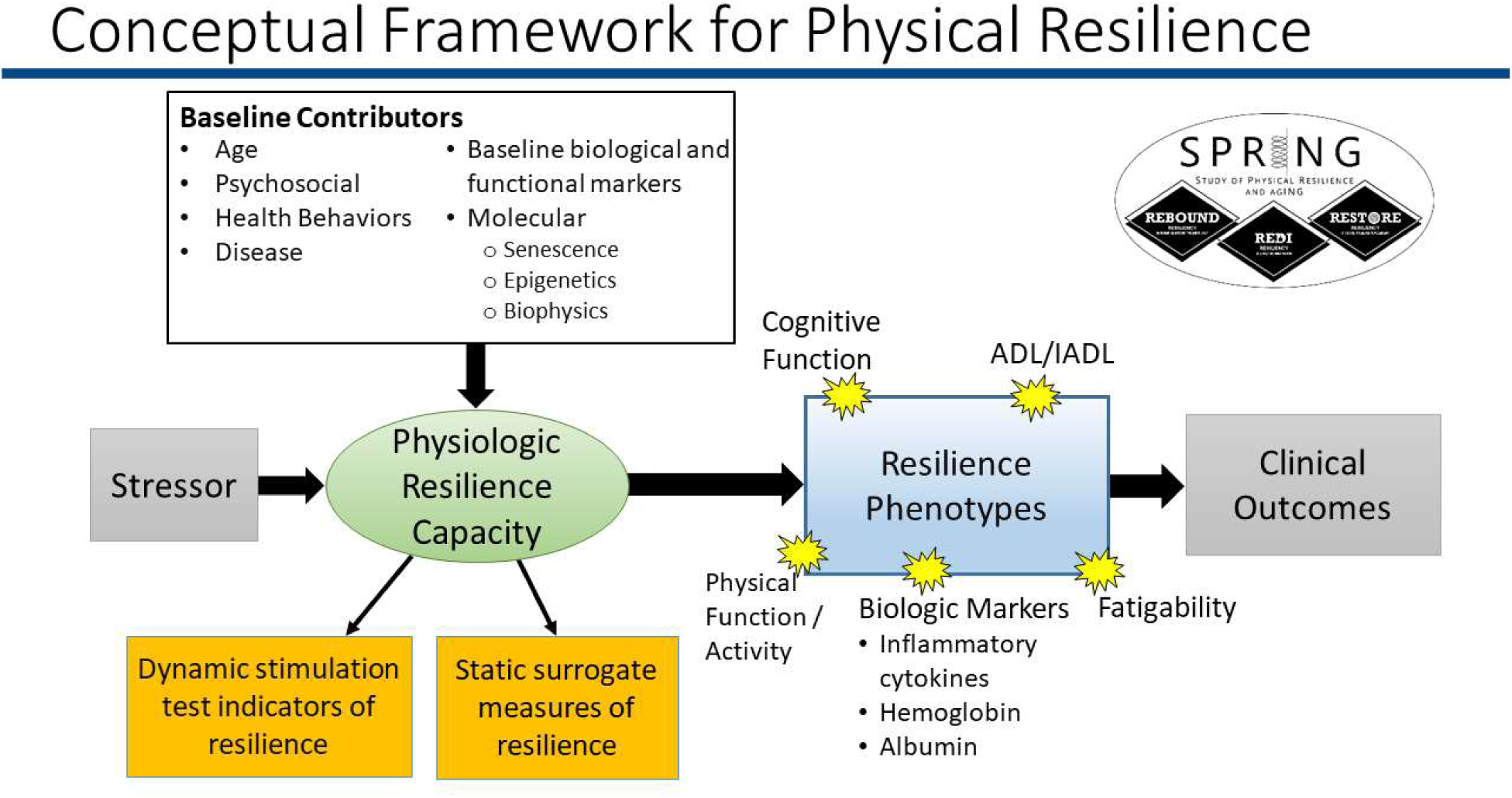
SPRING Conceptual Framework. Of primary focus in the current paper is the measurement of “physiologic resilience” using dynamic stimulation measures. Associations of such a measurement with potential determinants of resilience and static, “surrogate” measures of physiologic resilience also are evaluated in the present paper. The figure additionally illustrates that clinical stressors to be experienced—such as those anchoring SPRING—operate in the context of the physiologic resilience capacity, and the capacity itself is a product of one’s physical characteristics and health as well as (not shown) the social and environmental milieu in which one is embedded. “Physical resilience” refers to observed trajectories of phenotypic measures over the time period shortly preceding and then following the stressor experience both short and longer term, and outcomes are the clinical results the stressors aim to achieve— survival, certainly, but also absence of adverse events, success in restoring function or eliminating disease, or the like. Permission to use Figure 1 from *Walston J, Varadhan R, Xue Q-L, et al. A Study of Physical Resilience and Aging (SPRING): Conceptual framework, rationale, and study design. J Am Geriatr Soc. 2023; 71*(*8*)*:2393-2405.* doi:10.1111/jgs.18483 was granted by the publisher John Wiley & Sons via the Copyright Clearance Center’s RightsLink® service.

**eTable 1.**
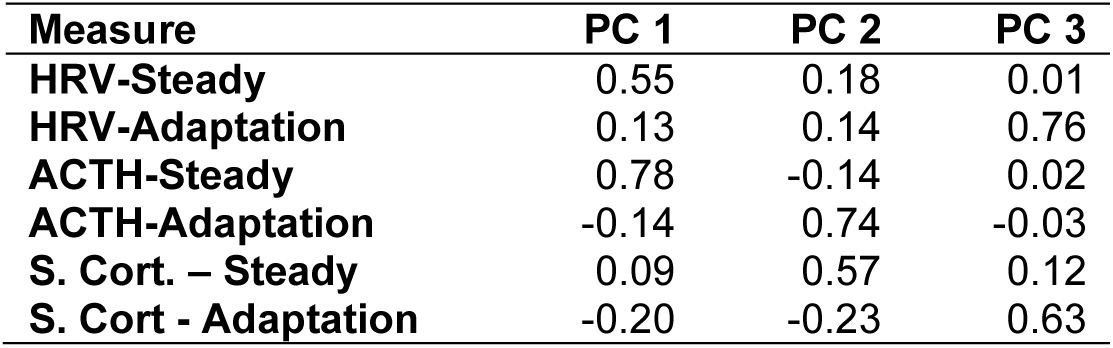
Varimax-rotated PCs loadings for 3-component solution, physiologic regulation metrics in SPRING. Complete case analysis (n=44).

**eFigure 2.**
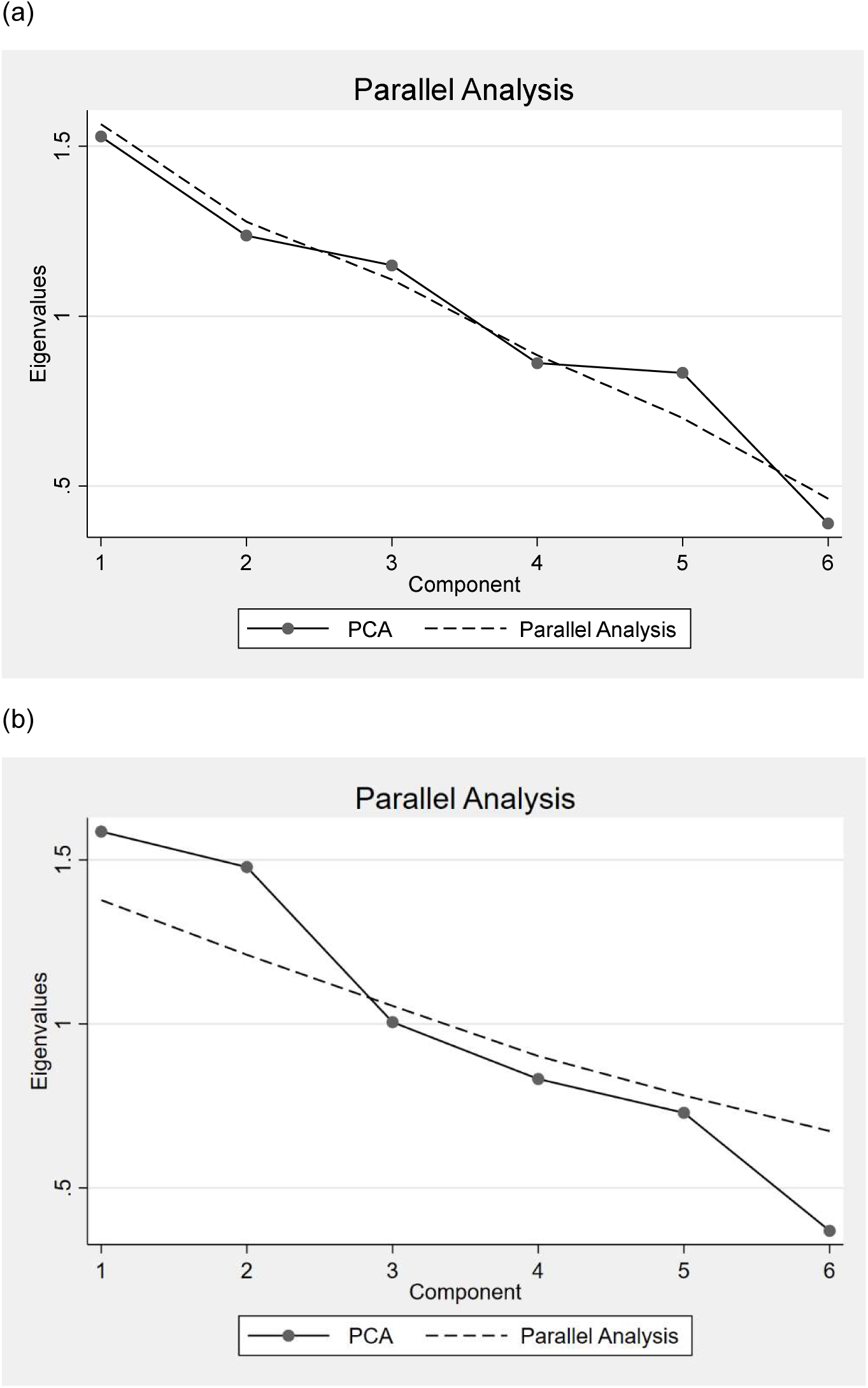
Parallel analysis plots, PCAs of (a) complete case data (n=44) and (b) PCA on average for each of 6 metrics over 10 multiple imputations (n=79). Eigenvalues for principal components analysis of 6 physiological functioning metrics measured in SPRING (solid line) are overlaid with eigenvalues from a principal components analysis of 6 randomly generated, independent variables (10 replicates; dashed line connects these). Dimensions of shared covariation are evidenced for observed data eigenvalues trending above the independently generated data eigenvalues: In (a) no dimensions of shared covariation are clearly evidenced, whereas in (b), 2 dimensions are evidenced.

### eMethods. Report on imputation-specific analyses

Parallel analysis applied to one replicate at a time after multiple imputation most often suggested 2 components (5 of 10 replicates), followed by 3 components (3 replicates) and 1 component (2 replicates).

In the eight cases in which 2 or 3 components were indicated, dimensions involving multi-measure steady state, adaptation, or their contrast were identified (Supplement Table 2). <u>When the first 2 components were rotated for</u> <u>these eight cases</u>, a component highly loading on the *HRV* and *ACTH* <u>steady state</u> measures was present in 7 of the 8 replicates; the remaining replicates additionally produced loadings of comparable magnitude for the *HRV* <u>adaptation</u> measure and the *negated ACTH* <u>adaptation</u> measure. The remaining component reliably loaded on *salivary cortisol* <u>adaptation</u>. In 5 of the replicates it also highly loaded on the *HRV* <u>adaptation</u>; in the remaining replicates, it contrasted the two *salivary cortisol* measures (once), or loadings for all remaining variables were small (twice). <u>When the first 3 components were rotated for these eight cases</u>, a component highly loaded on the *HRV* and *salivary cortisol* <u>adaptation</u> measures appeared in all cases, and in only one replicate did this component load appreciably on any other measure (*negated salivary cortisol* steady state). A second component highly loading on the *HRV* and *ACTH* <u>steady state</u> measures appeared in 7 out of the 8 replicates; *salivary cortisol* steady state also was highly loaded in one of these, and *negated ACTH adaptation* in one other. The remaining component was considerably variable but reliably involved *salivary cortisol* <u>steady state</u> or *ACTH* <u>adaptation</u>. In 2 replicates the two measures were contrasted (2 replicates); for remaining replicates, the component highly loaded on only *ACTH* <u>adaptation</u> (4 times) or only *salivary cortisol* <u>steady state (</u>twice).

In one of the two <u>cases with a single component indicated</u>, the leading component was highly loaded on the *HRV* and *salivary cortisol* <u>adaptation</u> measures. In the other, the overall pattern was to contrast the <u>steady state</u> measures *versus* the <u>adaptation</u> measures, with particularly high loadings for the salivary cortisol and ACTH adaptation measures (negative signs) and the salivary cortisol steady state measure. A summary of loading estimates for these two cases appears just below:

| Measure | Loading |  |
| --- | --- | --- |
|  | Case 1 | Case 2 |
| **HRV-Steady** | 0.07 | 0.15 |
| **HRV-Adaptation** | 0.61 | -0.32 |
| **ACTH-Steady** | 0.33 | 0.23 |
| **ACTH-Adaptation** | -0.19 | 0.41 |
| **S. Cort. – Steady** | -0.32 | 0.55 |
| **S. Cort - Adaptation** | 0.61 | -0.59 |

In the main manuscript, we synthesized information across imputations by performing PCA on the mean of each stimulus-response metric over replicates. We compared this approach with a second synthesis method: Performing PCA on the averages of the between-metric correlations (6-by-6 matrix) over replicates. In each case, PC scores were computed by applying the resulting loadings to the z-transformed metric values. Correlations between scores for the two methods exceeded 0.997 for both PCs.

**eTable 2.** Varimax-rotated PCs loadings, physiologic regulation metrics in SPRING, per multiply imputed replicate.

| Replicate | Measure | #Components | 2 COMP-PC1 | 2 COMP-PC2 | 3 COMP-PC1 | 3 COMP-PC2 | 3 COMP-PC3 |
| --- | --- | --- | --- | --- | --- | --- | --- |
| 1 | HRV – Steady State | 2 | 0.14 | 0.54 | 0.04 | 0.71 | -0.23 |
|  | HRV – Adaptation |  | 0.53 | 0.32 | 0.67 | 0.11 | 0.28 |
|  | ACTH – Steady State |  | -0.05 | 0.68 | -0.01 | 0.67 | 0.18 |
|  | ACTH – Adaptation |  | -0.28 | 0.34 | 0.04 | -0.04 | 0.86 |
|  | S. Cort. – Steady State |  | -0.50 | 0.18 | -0.43 | 0.13 | 0.28 |
|  | S. Cort. - Adaptation |  | 0.61 | -0.04 | 0.60 | -0.06 | -0.17 |
| 2 | HRV – Steady State | 3 | 0.09 | -0.09 | -0.06 | -0.18 | 0.70 |
|  | HRV – Adaptation |  | 0.13 | 0.76 | 0.16 | 0.76 | 0.02 |
|  | ACTH – Steady State |  | 0.18 | 0.29 | 0.05 | 0.20 | 0.71 |
|  | ACTH – Adaptation |  | 0.64 | 0.17 | 0.67 | 0.17 | -0.05 |
|  | S. Cort. – Steady State |  | 0.61 | -0.06 | 0.61 | -0.07 | 0.04 |
|  | S. Cort. - Adaptation |  | -0.40 | 0.55 | -0.38 | 0.56 | -0.02 |
| 3 | HRV – Steady State | 2 | 0.46 | -0.09 | 0.62 | -0.08 | -0.28 |
|  | HRV – Adaptation |  | 0.51 | 0.41 | 0.17 | 0.73 | 0.23 |
|  | ACTH – Steady State |  | 0.55 | 0.04 | 0.54 | 0.20 | -0.05 |
|  | ACTH – Adaptation |  | 0.46 | -0.36 | 0.47 | -0.05 | 0.35 |
|  | S. Cort. – Steady State |  | 0.08 | -0.38 | -0.09 | 0.03 | 0.81 |
|  | S. Cort. - Adaptation |  | 0.02 | 0.74 | -0.24 | 0.65 | -0.29 |
| 4 | HRV – Steady State | 2 | -0.07 | 0.63 | 0.66 | -0.06 | 0.01 |
|  | HRV – Adaptation |  | 0.60 | 0.23 | 0.08 | 0.72 | 0.23 |
|  | ACTH – Steady State |  | 0.31 | 0.55 | 0.58 | 0.30 | -0.11 |
|  | ACTH – Adaptation |  | -0.25 | 0.24 | -0.02 | 0.05 | 0.92 |
|  | S. Cort. – Steady State |  | -0.27 | 0.41 | 0.45 | -0.27 | 0.03 |
|  | S. Cort. - Adaptation |  | 0.63 | -0.14 | -0.13 | 0.56 | -0.31 |
| 6<sup>a</sup> | HRV – Steady State | 3 | -0.02 | 0.53 | -0.15 | 0.68 | -0.08 |
|  | HRV – Adaptation |  | 0.67 | 0.13 | 0.70 | 0.10 | 0.09 |
|  | ACTH – Steady State |  | 0.30 | 0.57 | 0.21 | 0.67 | 0.01 |
|  | ACTH – Adaptation |  | -0.12 | 0.33 | 0.09 | -0.12 | 0.79 |
|  | S. Cort. – Steady State |  | -0.26 | 0.50 | -0.16 | 0.21 | 0.59 |
|  | S. Cort. - Adaptation |  | 0.62 | -0.13 | 0.64 | -0.10 | -0.07 |
| 7 | HRV – Steady State | 2 | 0.01 | 0.59 | 0.20 | 0.54 | 0.24 |
|  | HRV – Adaptation |  | 0.59 | 0.26 | 0.65 | -0.02 | 0.07 |
|  | ACTH – Steady State |  | 0.15 | 0.57 | 0.36 | 0.13 | 0.56 |
|  | ACTH – Adaptation |  | -0.23 | 0.42 | -0.11 | 0.80 | -0.14 |
|  | S. Cort. – Steady State |  | -0.35 | 0.29 | -0.19 | -0.17 | 0.73 |
|  | S. Cort. - Adaptation |  | 0.68 | -0.07 | 0.61 | -0.14 | -0.29 |
| 8 | HRV – Steady State | 2 | 0.58 | -0.09 | -0.08 | 0.71 | 0.01 |
|  | HRV – Adaptation |  | 0.38 | 0.44 | 0.67 | 0.10 | 0.18 |
|  | ACTH – Steady State |  | 0.60 | 0.09 | 0.10 | 0.69 | -0.04 |
|  | ACTH – Adaptation |  | 0.34 | -0.37 | 0.11 | -0.03 | 0.82 |
|  | S. Cort. – Steady State |  | 0.16 | -0.55 | -0.29 | 0.04 | 0.54 |
|  | S. Cort. - Adaptation |  | 0.09 | 0.59 | 0.66 | -0.10 | -0.08 |
| 9<sup>a</sup> | HRV – Steady State | 3 | 0.63 | -0.07 | 0.71 | -0.05 | -0.04 |
|  | HRV – Adaptation |  | 0.34 | 0.45 | 0.10 | 0.65 | 0.38 |
|  | ACTH – Steady State |  | 0.61 | 0.09 | 0.65 | 0.11 | -0.05 |
|  | ACTH – Adaptation |  | 0.28 | -0.47 | 0.23 | -0.34 | 0.39 |
|  | S. Cort. – Steady State |  | 0.20 | -0.27 | -0.08 | 0.03 | 0.81 |
|  | S. Cort. - Adaptation |  | 0.03 | 0.70 | -0.03 | 0.67 | -0.21 |

Note. Data shown for 8 out of 10 replicates for which 2 or 3 components were indicated by parallel analysis (n=79 per replicate).
Note. HRV = heart rate variability; ACTH = adrenal corticotropic hormone test; S. Cort. = salivary cortisol
a In replicates 5 and 10, parallel analysis indicated a single component. Component loadings for these are described in the eMethods.

**eTable 3.**
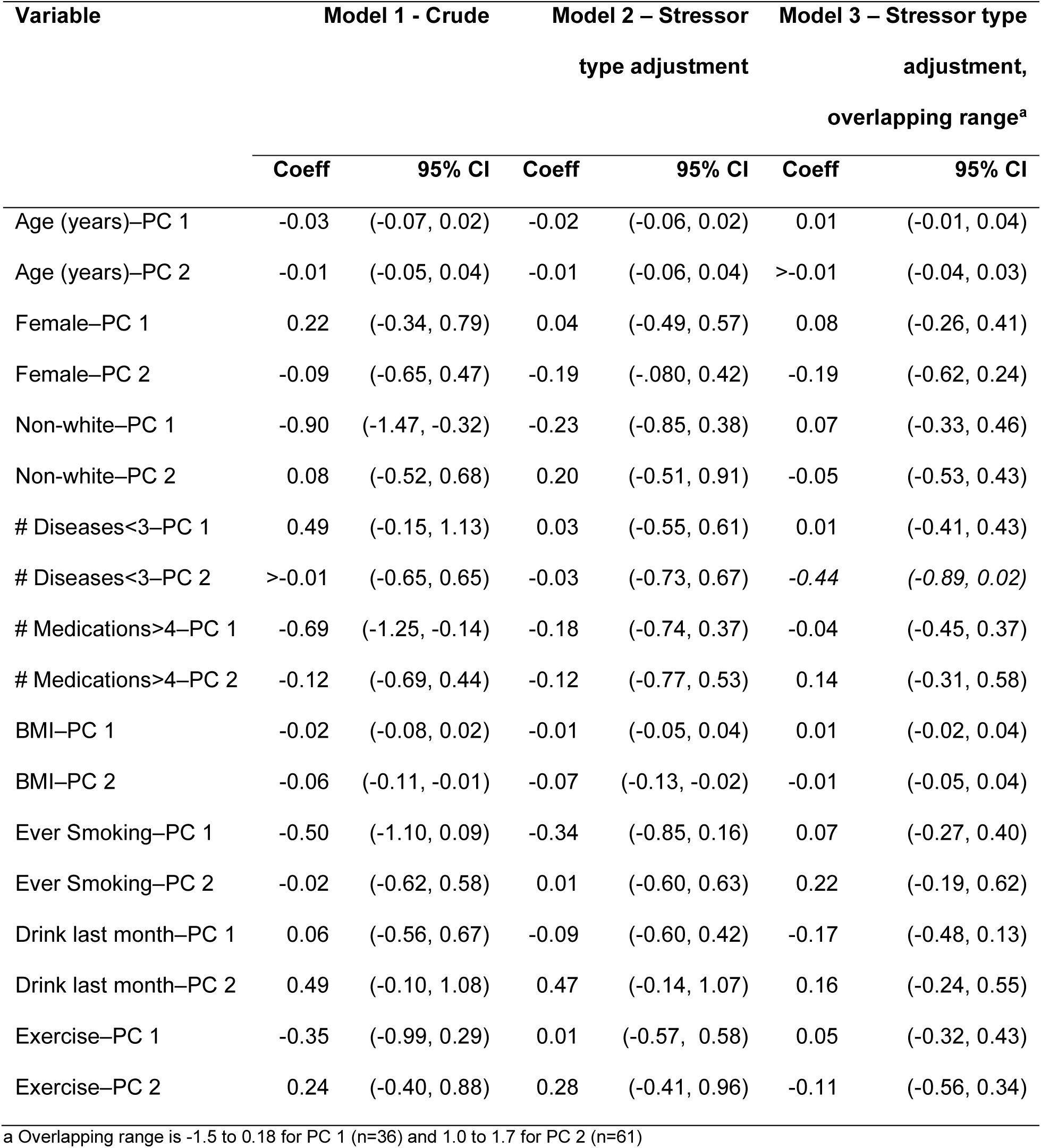
Linear regressions of multiply imputed mean principal component (PC) scores on potential determinants of resilience in SPRING Pilot, with stressor type adjustment.

| Variable | Model 1 - Crude |  | Model 2 – Stressor type adjustment |  | Model 3 – Stressor type adjustment, overlapping range<sup>a</sup> |  |
| --- | --- | --- | --- | --- | --- | --- |
|  | Coeff | 95% CI | Coeff | 95% CI | Coeff | 95% CI |
| Age (years)–PC 1 | -0.03 | (-0.07, 0.02) | -0.02 | (-0.06, 0.02) | 0.01 | (-0.01, 0.04) |
| Age (years)–PC 2 | -0.01 | (-0.05, 0.04) | -0.01 | (-0.06, 0.04) | >-0.01 | (-0.04, 0.03) |
| Female–PC 1 | 0.22 | (-0.34, 0.79) | 0.04 | (-0.49, 0.57) | 0.08 | (-0.26, 0.41) |
| Female–PC 2 | -0.09 | (-0.65, 0.47) | -0.19 | (-0.80, 0.42) | -0.19 | (-0.62, 0.24) |
| Non-white–PC 1 | -0.90 | (-1.47, -0.32) | -0.23 | (-0.85, 0.38) | 0.07 | (-0.33, 0.46) |
| Non-white–PC 2 | 0.08 | (-0.52, 0.68) | 0.20 | (-0.51, 0.91) | -0.05 | (-0.53, 0.43) |
| # Diseases<3–PC 1 | 0.49 | (-0.15, 1.13) | 0.03 | (-0.55, 0.61) | 0.01 | (-0.41, 0.43) |
| # Diseases<3–PC 2 | >-0.01 | (-0.65, 0.65) | -0.03 | (-0.73, 0.67) | -0.44 | (-0.89, 0.02) |
| # Medications>4–PC 1 | -0.69 | (-1.25, -0.14) | -0.18 | (-0.74, 0.37) | -0.04 | (-0.45, 0.37) |
| # Medications>4–PC 2 | -0.12 | (-0.69, 0.44) | -0.12 | (-0.77, 0.53) | 0.14 | (-0.31, 0.58) |
| BMI–PC 1 | -0.02 | (-0.08, 0.02) | -0.01 | (-0.05, 0.04) | 0.01 | (-0.02, 0.04) |
| BMI–PC 2 | -0.06 | (-0.11, -0.01) | -0.07 | (-0.13, -0.02) | -0.01 | (-0.05, 0.04) |
| Ever Smoking–PC 1 | -0.50 | (-1.10, 0.09) | -0.34 | (-0.85, 0.16) | 0.07 | (-0.27, 0.40) |
| Ever Smoking–PC 2 | -0.02 | (-0.62, 0.58) | 0.01 | (-0.60, 0.63) | 0.22 | (-0.19, 0.62) |
| Drink last month–PC 1 | 0.06 | (-0.56, 0.67) | -0.09 | (-0.60, 0.42) | -0.17 | (-0.48, 0.13) |
| Drink last month–PC 2 | 0.49 | (-0.10, 1.08) | 0.47 | (-0.14, 1.07) | 0.16 | (-0.24, 0.55) |
| Exercise–PC 1 | -0.35 | (-0.99, 0.29) | 0.01 | (-0.57, 0.58) | 0.05 | (-0.32, 0.43) |
| Exercise–PC 2 | 0.24 | (-0.40, 0.88) | 0.28 | (-0.41, 0.96) | -0.11 | (-0.56, 0.34) |

a Overlapping range is -1.5 to 0.18 for PC 1 (n=36) and 1.0 to 1.7 for PC 2 (n=61)

